# COVID-19 Vaccine Uptake among U.S. Child Care Providers

**DOI:** 10.1101/2021.07.30.21261383

**Authors:** Kavin M. Patel, Amyn A. Malik, Aiden Lee, Madeline Klotz, John Eric Humphries, Thomas Murray, David Wilkinson, Mehr Shafiq, Inci Yildirim, Jad A. Elharake, Rachel Diaz, Chin Reyes, Saad B. Omer, Walter S. Gilliam

**Affiliations:** Yale School of Medicine, New Haven, CT; Yale Institute for Global Health, New Haven, CT; Tobin Center for Economic Policy, Yale University, New Haven, CT; Department of Economics, Yale University, New Haven, CT; Department of Epidemiology of Microbial Diseases, Yale School of Public Health, New Haven, CT; Department of Pediatrics, Yale School of Medicine, New Haven, CT; Mailman School of Public Health, Columbia University, New York, NY; Yale Child Study Center, Yale School of Medicine, New Haven, CT; Yale School of Public Health, New Haven, CT; Yale School of Nursing, New Haven, CT

## Abstract

**Objectives:** Ensuring a high COVID-19 vaccine uptake among U.S. child care providers is crucial to mitigating the public health implications of child-to-staff and staff-to-child transmission of SARS-CoV-2; however, the vaccination rate among this group is unknown.

**Methods:** To characterize the vaccine uptake among U.S. child care providers, we conducted a cross-sectional survey of the child care workforce. Providers were identified through various national databases and state registries. A link to the survey was sent via email between May 26 and June 23, 2021. Out of 44,771 potential respondents, 21,663 responded (48.4%).

**Results:** Overall COVID-19 vaccine uptake among U.S. child care providers (78.1%, 95% CI [77.3% to 78.9%]) was higher than that of the U.S. adult population (65%). Vaccination rates varied from 53.5% to 89.4% between states. Vaccine uptake differed significantly (p < .01) based on respondent age (70.0% for ages 25-34, 91.5% for ages 75-84), race (70.0% for Black or African Americans, 92.5% for Asian-Americans), annual household income (70.7% for <$35,000, 85.0% for>$75,000), and childcare setting (72.9% for home-based, 79.7% for center-based).

**Conclusions:** COVID-19 vaccine uptake among U.S. child care providers was higher than that of the general U.S. adult population. Those who were younger, lower income, Black or African American, resided in states either in the Mountain West or the South, and/or worked in home-based childcare programs reported the lowest rates of vaccination; state public health leaders and lawmakers should prioritize these subgroups for placement on the policy agenda to realize the largest gains in vaccine uptake among providers.

**Tables of Contents Summary:** This article describes the results of a national survey of childcare providers to determine the overall COVID-19 vaccine uptake and the gaps in vaccine coverage.

**What’s Known on This Subject:** Ensuring a high COVID-19 vaccine uptake among U.S. child care providers is crucial to mitigating the public health implications of child-to-staff and staff-to-child transmission of SARS-CoV-2; however, the vaccination rate among this group is unknown.

**What This Study Adds:** While the vaccine uptake among U.S. child care providers was higher than that of U.S. adults, certain subgroups continue to warrant focused attention for outreach and/or placement on the policy agenda.

## INTRODUCTION

Promoting coronavirus disease-2019 (COVID-19) vaccine uptake among U.S. child care providers is a vital step in restoring the U.S. to pre-pandemic functionality. Accurately billed as ‘the workforce behind the workforce,’^1^ child care providers supply an essential service to parents seeking to reenter the U.S. labor market.^2^ Evidence of this can be found in national surveys^3^ conducted by the Department of Education which estimated that almost 90 percent of parents in dual income households relied on some type of child care arrangement prior to the pandemic.^4^ Furthermore, in the aftermath of the pandemic, lack of availability of child care services and school closures have been among the leading causes preventing parents from returning to the workforce.^5^

In addition to the economic incentive, there exists a public health imperative to protect child care providers through vaccination given the potential implications of bidirectional transmission of SARS-CoV-2 between staff and children. Staff-to-child transmission may lead to COVID-19 among children, many of whom tend to be either asymptomatic or pauci-symptomatic relative to adults, and who may unwittingly transmit the infection to household contacts.^6^ Household contacts may include parents and grandparents, many of whom are elderly and/or have underlying medical conditions, which place them at higher risk for COVID-19-related morbidity and mortality.^7^ While the risk is low^8^, it is possible that child-to-staff transmission of SARS-CoV-2 may lead to COVID-19 among child care providers^8^, a disproportionate number of whom are members of racial and ethnic minority groups^9^ also at higher risk for COVID-19-related complications. Ensuring a high vaccine uptake among child care providers, thus, extends beyond the immediate issue of personal health and serves as a mechanism to promote economic recovery, public health, and health equity.

Despite the importance of ensuring a high vaccine uptake among U.S. child care providers, little is known about the group’s current vaccination status. To better characterize the current status of vaccine uptake among child care providers, we conducted a large-scale multistate cross-sectional survey of the U.S. child care workforce.

## METHODS

### Survey Administration

We conducted a cross-sectional survey via Qualtrics (Qualtrics, Provo, UT) of child care providers throughout the U.S. and territories. The survey was self-administered via email and available in both English and Spanish. A prenotification email was sent out through Yale University notifying potential respondents to expect an email from Qualtrics regarding the survey. The survey was active between May 26, 2021 and June 23, 2021; 11 reminders for survey completion were sent out spaced 2-3 days apart from each other. Child care providers contacted were those who had expressed an interest in participating in follow-up surveys from a prior survey conducted in May 2020 on COVID-19 transmission within U.S. child care programs.^8^ The original respondents were identified through two national child care provider databases and numerous state registries as follows: (1) Child Care Aware America; (2) National Association of Education for the Young Children; and (3) various state registries coordinated by the National Workforce Registry Alliance (out of a total of 41 states with child care provider registries, 28 states agreed to participate, 11 states were unable to secure permissions in time, and 2 states declined). Eligibility criteria for the survey included individuals who were employed as child care providers at any point during or just prior to the COVID-19 pandemic, either remotely or in-person, including both center-based childcare providers and home-based (family-based) child care providers. Respondents provided informed consent prior to data collection. Respondents were entered into a random drawing to be one of 20 selected to receive a $500 incentive. All respondents also were invited to participate in an optional free webinar where results of the study would be discussed. The research protocol was approved by the Yale University Institutional Review Board (IRB protocol number: 2000028232).

### Survey Content and Design

All questions used in the current analyses were close-ended questions with either nominal or ordinal answering scales. For questions with ordinal answering scales, positive and negative options were balanced. The survey was composed of questions designed to expand on the following characteristics of child care providers: demographics (i.e., age, race/ethnicity, and annual income level), current employment status within child care industry, history of COVID (i.e., previous positive COVID-19 test result and/or COVID-19 related hospitalization), vaccination status, vaccination date, vaccine brand, likelihood of vaccination if unvaccinated, reasons for non-vaccination pertaining to structural issues (e.g., lack of transportation, inability to get time off of work etc.) and vaccine hesitancy (e.g., vaccine-related concerns regarding lack of safety and/or efficacy, disease-related concerns regarding low perceived risk of disease acquisition and/or disease severity, etc.), child care type (i.e., center-based versus home-based), and leadership position (i.e., director or not director).

### Data Analysis

Descriptive statistics (frequencies, percentages) were calculated for the sample demographic characteristics. Additionally, the frequency and percentage of COVID-19 vaccine uptake was calculated at the national and state levels. States were determined based on the child care provider’s reported zip code. Data were weighted based on age, race, Hispanic origin, and state to match employed child care workers (occupation code: 4600) 18 years of age or older in the United States (50 states and Washington D.C.), based on the 2015-2019 American Community Survey (ACS). Weights were trimmed top and bottom at 2.5%. All reported percentages and data analytics correspond to weighted estimates unless otherwise specified. Pearson’s chi squared test was used for assessing the association between both the unweighted and weighted categorical variables. Data were analyzed using Stata version 16 (StataCorp, College Station, Texas).

## RESULTS

A total of 21,663 out of 44,771 to whom the survey was sent completed the survey yielding a response rate of 48.4%; of these, 20,013 (44.7% of those we sent the survey) respondents met the eligibility criteria for inclusion. Overall vaccine uptake among all eligible respondents was 78.2% (unweighted 77.7%), and vaccine uptake among those still actively working as child care providers at the time of survey completion (92% of respondents) was 78.4% versus 75.8% for those not currently working in child care (χ^2^ = 2.86; *p* = 0.09). Most respondents reported receiving either Pfizer-BioNTech (50.5%) or Moderna (42.1%) with a lesser proportion reporting receiving Johnson & Johnson (6.9%) or another vaccine (0.6%). The most commonly reported month for receiving the first dose was March 2021 (33.1%), with proportions steeply declining after March 2021.

### Vaccine Uptake by State

Vaccine uptake between states varied from a low of 53.5% in WY to a high of 89.4% in MA. The states with the highest vaccination rates tended to be in either the New England area (e.g., MA – 89.4%, NH – 88.0%; CT – 86.4%) or the Pacific West (e.g., WA – 85.4%, OR – 82.9%, CA – 81.8%) whereas the states with the lowest vaccination rates tended to be in either the South (e.g., OK – 62.4%, FL – 63.8%, AL – 67.7%) or the Mountain West (e.g., WY – 53.5%, MT – 64.4%, UT – 70.8%), although, the latter were limited by small sample sizes. State-by-state vaccine uptake among child care providers is reported in *Figure 1*.

**Figure 1:**
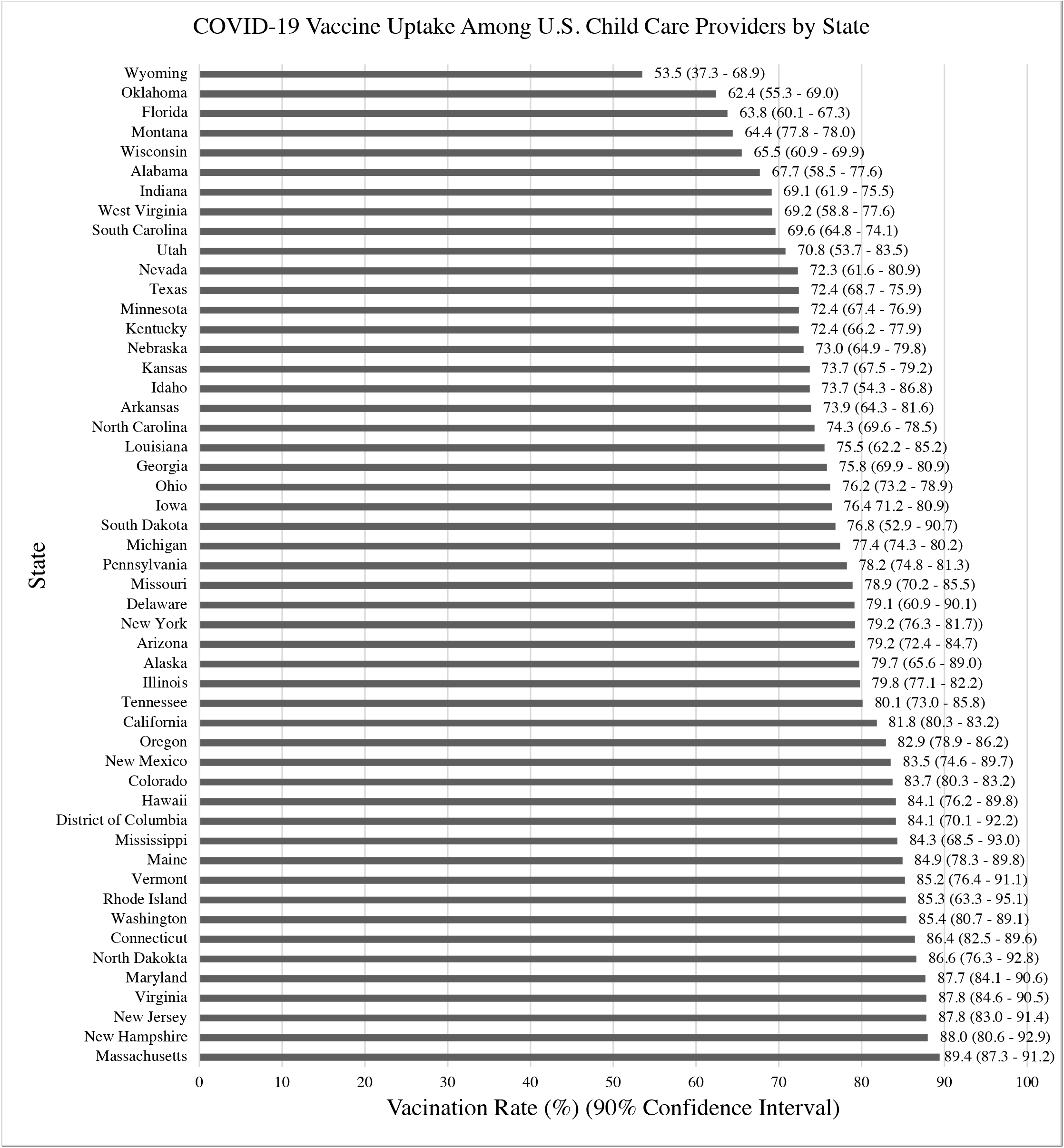
COVID-19 Vaccine Uptake Among U.S. Child Care Providers by State. Vaccine uptake was highest in the New England Area and the Pacific West, and lowest in the Mountain West and the South.

### Vaccine Uptake by Demographics

As shown in Table 1, vaccine uptake was higher among respondents who were older: the vaccination rate of those between 25-34 years of age was 70.0% whereas the vaccination rate of those between 75-84 years of age was 91.5% (χ^*2*^ *= 31*.*59; p < 0*.*01*), with progressively higher rates as age groups became older. Vaccine uptake was lower among Black or African American providers at 70.0% and higher among Asian Americans at 92.5% (χ^*2*^ *= 17*.*03; p < 0*.*01*). Annual household income level was associated with higher vaccine uptake with those earning <$35,000 having a vaccination rate of 70.7% and those earning >$75,000 having a vaccination rate of 85.0% (χ^*2*^ *= 52*.*47; p < 0*.*01*), with progressively higher rates as household income increases. A history of COVID-19 was associated with lower vaccine uptake at 70.5% versus 79.6% in those without a history of COVID-19 (χ^*2*^*= 63*.*46; p < 0*.*01*).

**Table 1:** Vaccination Rates of U.S. Child Care Providers by Selected Characteristics. The overall COVID-19 vaccine uptake among US child care providers was found to be higher than that of the general U.S. adult population: 78% versus 65% respectively. Those who were young, low income, Black or African American, and/or working in home-based child care centers reported the lowest rates of vaccination.

| Table 1: Vaccination Rates of Child Care Providers by Selected Characteristics |  |  |  |  |  |  |
| --- | --- | --- | --- | --- | --- | --- |
|  |  | Sample Size | Unweighted |  | Weighted |  |
|  |  |  | Percent | P value | Percent | P value |
| Overall |  |  |  |  |  |  |
| All Respondents |  | 20013 | 80.2 |  | 78.1 |  |
| Age Group |  |  |  |  |  |  |
| 18 - 24 | | 380 | 73.4 | $\chi^2 = 461.10$ | 73.0 | $\chi^2 = 31.59$ |
| 25 - 34 | | 2400 | 70.2 | $p < 0.01$ | 70.0 | $p < 0.01$ |
| 35 - 44 |  | 4637 | 75.3 |  | 74.4 |  |
| 45 - 54 |  | 6053 | 80.7 |  | 80.2 |  |
| 55 - 64 |  | 5078 | 86.6 |  | 86.3 |  |
| 65 - 74 |  | 1339 | 90.2 |  | 88.7 |  |
| 75 - 84 |  | 94 | 92.5 |  | 91.5 |  |
| Race |  |  |  |  |  |  |
| White | | 14848 | 81.6 | $\chi^2 = 209.67$ | 79.8 | $\chi^2 = 17.03$ |
| Black or African American | | 2132 | 71.2 | $p < 0.01$ | 70.0 | $p < 0.01$ |
| American Indian or Alaskan Native |  | 172 | 76.1 |  | 77.9 |  |
| Asian |  | 567 | 92.7 |  | 92.5 |  |
| Native Hawaiian or Other Pacific Islander |  | 53 | 81.1 |  | 79.9 |  |
| Multiracial |  | 409 | 71.3 |  | 71.3 |  |
| Prefer not to answer |  | 1278 | 78.7 |  | 76.6 |  |
| Ethnicity |  |  |  |  |  |  |
| Hispanic | | 3257 | 81.1 | $\chi^2 = 1.47$ | 78.7 | $\chi^2 = .27$ |
| Not Hispanic | | 16377 | 80.2 | $p = 0.22$ | 78.1 | $p = 0.60$ |
| Annual Household Income |  |  |  |  |  |  |
| <\$35,000 | | 3499 | 71.8 | $\chi^2 = 335.55$ | 70.7 | $\chi^2 = 52.47$ |
| \$35,000 - \$49,999 | | 3308 | 77.1 | $p < 0.01$ | 75.4 | $p < 0.01$ |
| \$50,000 - \$74,999 | | 4151 | 80.3 | | 78.2 | |
| >\$75,000 | | 6466 | 86.4 | | 85.0 | |
| History of COVID-19 |  |  |  |  |  |  |
| Yes | | 2879 | 71.2 | $\chi^2 = 173$ | 70.5 | $\chi^2 = 63.46$ |
| No | | 16982 | 81.8 | $p < 0.01$ | 79.6 | $p < 0.01$ |
| Setting of Child Care Provider |  |  |  |  |  |  |
| Home-based | | 5112 | 75.6 | $\chi^2 = 83.51$ | 72.9 | $\chi^2 = 50.92$ |
| Center-based | | 12887 | 81.7 | $p < 0.01$ | 79.7 | $p < 0.01$ |
| Types of Center-Based Settings |  |  |  |  |  |  |
| For-Profit | | 4160 | 79.9 | $\chi^2 = 96.89$ | 78.5 | $\chi^2 = 7.78$ |
| Not-For-Profit | | 3101 | 86.3 | $p < 0.01$ | 84.1 | $p < 0.01$ |
| School-Based |  | 1706 | 82.8 |  | 81.1 |  |
| Head Start or Early Head Start |  | 1456 | 75.6 |  | 72.9 |  |
| Drop-in |  | 51 | 72.5 |  | 63.4 |  |
| Faith Based |  | 1744 | 81.3 |  | 79.6 |  |
| Other |  | 650 | 84.1 |  | 82.5 |  |
| Director of Child Care Program |  |  |  |  |  |  |
| Yes | | 18402 | 80.4 | $\chi^2 = 5.52$ | 78.4 | $\chi^2 = 2.8$ |
| No | | 1575 | 78.0 | $p < 0.01$ | 75.8 | $p < 0.09$ |

### Vaccine Uptake by Setting of Child Care

Vaccine uptake was higher among center-based child care providers relative to home-based child care providers with reported vaccination rates of 79.7% and 72.9% respectively (χ^*2*^ *= 50*.*92; p < 0*.*01*). Notably, racial minority providers comprise a higher proportion of home-based childcare settings relative to center-based childcare settings. Among the different types of center-based child care programs, vaccine uptake was as follows: for profit centers – 78.5%, not-for profit agency centers – 84.1%, school-based – 81.1%, Head Start or Early Head Start (federally funded child care centers) – 72.9%, drop-in child care (the children change every day) – 63.4%, faith-based child care programs – 79.6% and other center-based – 82.5% (χ^*2*^ *= 7*.*78; p < 0*.*01*).

### Reasons for Nonvaccination

Among those who were unvaccinated, only 5.0% were “absolutely certain” that they would get vaccinated in the future, 6.9% were “very likely,” 28.2% were “somewhat likely,” and 59.2% were “not likely” (0.4% did not respond). Among those who reported being only “somewhat likely” or “not likely” to vaccinate in the future, the most common reasons for nonvaccination included concerns about vaccine safety (79.9%), concerns about the speed of vaccine development (66.8%), and lack of trust in the vaccine development process (32.0%). The most common structural barriers to vaccination were inability to take time off work (5.4%), too busy for vaccination (3.5%), and difficulty with scheduling a vaccine appointment date (2.7%). A complete list of the reported reasons for nonvaccination among respondents can be found in *Figure 2*.

**Figure 2:**
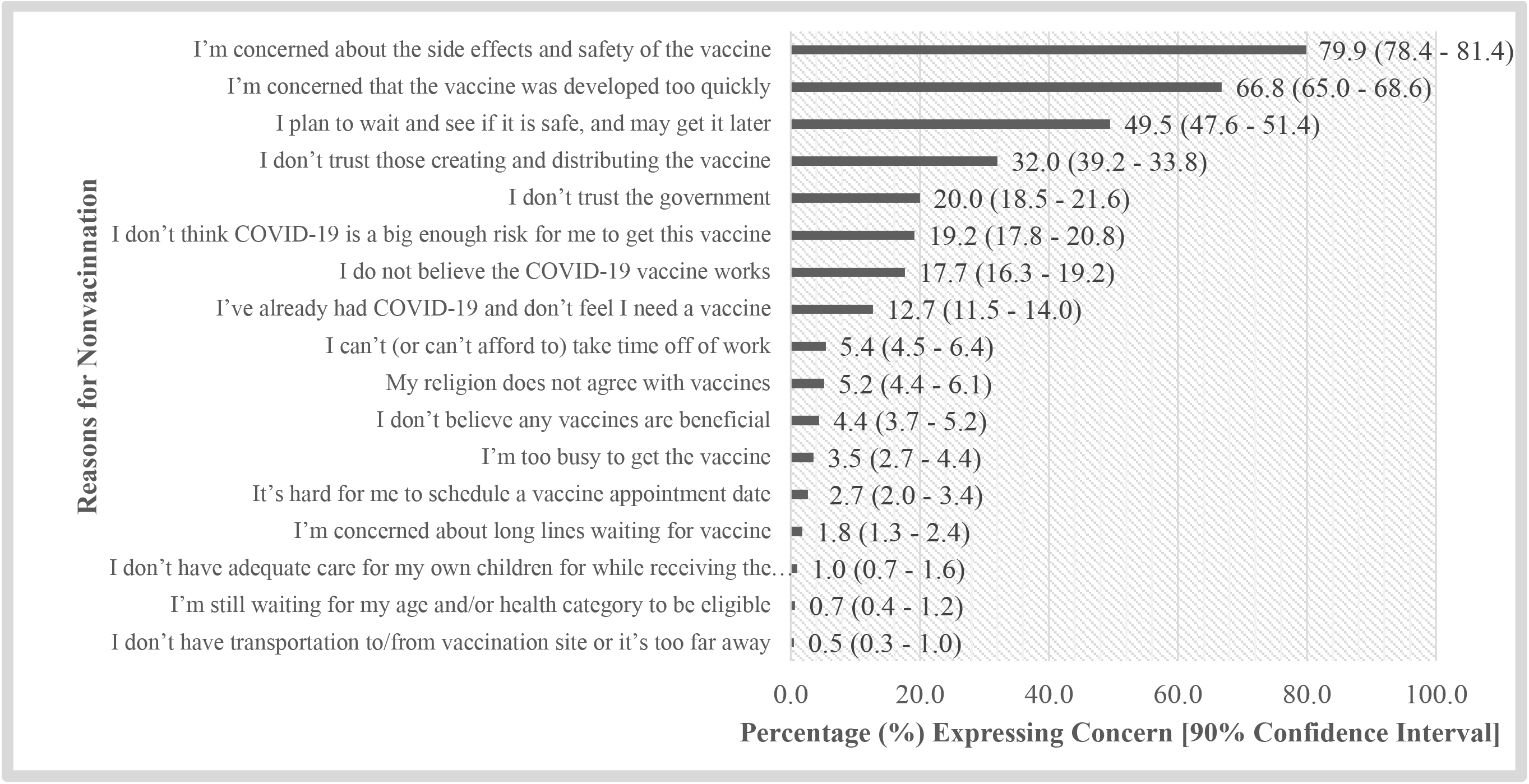
Reasons for Nonvaccination Among U.S. Child Care Providers. The most commonly reported reasons for nonvaccination among child care providers had to do with vaccine hesitancy rather than structural barriers to vaccination.

## DISCUSSION

Overall vaccine uptake among U.S. child care providers (78.2% (95% CI: 77.3 to 78.9)) was significantly higher than that of the general U.S. adult population (65%) at the time of the survey.^10^ Notably, the vaccination rate among those child care providers actively working within the child care industry (92% of respondents) at the time of the survey was even higher (78.4%). Of those reporting having not yet received the COVID-19 vaccine, another 11.9% stated that they were “absolutely certain” (5.0%) or “very likely” (6.9%) to get vaccinated in the future, suggesting that the final vaccine uptake among child care providers may settle around 90%. Our results are similar to those described in an April 6, 2021 media report^11^ released by the Center for Disease Control and Prevention based on unpublished data. Additionally, our study provides a detailed breakdown of the vaccine uptake among child care providers based on demographics, location and setting of child care.

The increased vaccine uptake among child care providers relative to the general population may be explained by an increased disease salience among this group pertaining to COVID-19. As an example, congregate settings (e.g., schools^12^, child care programs^6^, nursing homes^13^, prisons^14^) without appropriate infection mitigation measures are known vectors for community transmission. Also, young children were ineligible for pharmaceutical interventions such as vaccination at the time of data collection and may be unable to effectively adhere to nonpharmaceutical behavioral interventions. This is particularly relevant to child care providers who, unlike K-12 teachers, care for children who are mostly under five years old.^8^ Child care providers may also feel it is their responsibility to protect the currently unvaccinated children in their care. Furthermore, while no state currently requires child care providers to be vaccinated, some child care agencies may require vaccination as a condition of employment.

While overall vaccine uptake among U.S. child care providers was significantly higher than that of the general U.S. adult population, the pattern of relative differences in vaccination rates were similar in terms of demographics, state where they provided care, and reasons for non-vaccination.^15,16^ Those who were younger, lower income and/or Black or African-American reported the lowest rates of vaccination; conversely, those who were elderly, higher income and Asian-American reported the highest rates of vaccination (one exception is child care providers between 18 and 24 years of age who reported higher vaccine uptake relative to the 25-34 year old group, perhaps because it was required by some higher educational institutions as a condition of enrollment^17^). Vaccine uptake was found to be lowest in the Mountain West and the South, and highest in the New England area and the Pacific West. The most commonly cited reasons for nonvaccination were related to vaccine hesitancy rather than structural issues related to a lack of access; these included concerns about vaccine safety, lack of trust in the vaccine development process and/or lack of trust in the government among other reasons.

Finally, COVID-19 vaccine uptake varied among the different child care settings. Center-based childcare providers reported a higher vaccination rate compared to home-based child care providers (79.7% and 72.9%, respectively). One reason for this may be that home-based child providers skewed more heavily Black or African American, Native American and/or Hispanic (although the latter was not significant) relative to center-based child care providers; minority groups have been known to have lower vaccine uptake relative to non-Hispanic Whites.^18^ Another consideration that could account for the difference may be increased disease salience among center-based over home-based child care providers arising from variation in group sizes and the potential that some child care employers may have required vaccination of providers as a condition for returning to work. Both group size and child-to-staff ratios among child care settings are tightly regulated^19^, with home-based child care settings having a lower cap on group size relative to center-based child care settings. For example, in Connecticut^20^, licensed center-based child care centers must have more than 12 children; whereas home-based child care settings in a private family residence can have no more than 6. It is thus possible that center-based child care providers may have had a higher vaccination rate compared to their home-based counterparts given the increased perception of disease acquisition arising from the higher number of children and staff with whom they would come into contact, even though person density may be higher in home-based settings.

Limitations to our study include the following. First, our sample sizes from some states (mostly in the Mountain West) were relatively low and with a sampling error of above 5%. Second, the respondents of our survey were those who, last year, had completed our previous survey, indicated a willingness to complete later surveys, and then completed a one-year follow up; therefore, it’s possible that this subsample of respondents who are reliable survey takers may differ from the population in meaningful ways that could bias generalization. The major strengths to our study include a large national sample weighted to representativeness and a strong response rate.

## CONCLUSION

COVID-19 vaccine uptake among U.S. child care providers was found to be higher than that of the general U.S. adult population. The lowest vaccination rate was reported among child care providers who were younger, lower income, Black or African-American, resided in states either in the Mountain West or the South, and/or worked in home-based child care centers. State public health leaders and lawmakers should prioritize these subgroups for outreach and placement on the policy agenda to realize the largest possible gains in vaccine uptake among child care providers. Efforts to promote COVID-19 vaccine uptake among child care providers take on added significance when considering the emergence of the more transmissible Delta Variant, which in July 2021 became dominant stain of SARS-CoV-2 in the U.S.^21^ Children under the age of 12 remain ineligible for COVID-19 vaccination, and as such, the limiting factor in ensuring an adequate supply of child care services will be contingent on protecting the personal health of child care providers.

## Data Availability

N/A

## Abbreviations

None

## Acknowledgements

This study was supported by The Andrew & Julie Klingenstein Family Fund, Esther A. & Joseph Klingenstein Fund, Heising-Simons Foundation, W.K. Kellogg Foundation, Foundation for Child Development, Early Educator Investment Collaborative, Scholastic Inc, Yale Institute for Global Health, and Tobin Center for Economic Policy at Yale University. Invaluable assistance with obtaining child care provider contact information was provided by the National Workforce Registry Alliance (and its network of state child care workforce registries), Child Care Aware of America, and National Association for the Education of Young Children. Drs. Amalia Londono Tobon and Adrián Cerezo Caballero provided Spanish translations and back translations of the survey measures and recruitment information. Alicia Alonso, Catherine Chang, Renee Dauerman, Stella FitzGerald, Harleen Kaur, Emma Knight, and Helen Mooney provided assistance in qualitative data categorization of respondent comments.

